# Bidirectional relationships between mental health problems and urinary incontinence in women: a two-sample Mendelian randomization analysis

**DOI:** 10.64898/2026.02.06.26345734

**Authors:** Rochelle Knight, Ana Goncalves Soares, Kimberley Burrows, Abigail Fraser, Tom M Palmer, Rufus Cartwright, Carol Joinson

## Abstract

**Objective:** Comorbidity between urinary incontinence (UI) and affective disorders, including anxiety and depression, is well established in cross-sectional studies and prospective bidirectional associations have also been reported. It is, however, unclear whether these associations are causal. We applied two-sample Mendelian randomization (MR) analysis to examine if there are causal bidirectional relationships between UI and anxiety, depression, and neuroticism in women.

**Materials and methods:** We used summary level data from independent genome-wide association studies (GWAS) to estimate the bidirectional causal effects of UI and its subtypes (stress urinary incontinence (SUI), urgency urinary incontinence (UUI), and mixed urinary incontinence (MUI), n=10,931) on anxiety (n=83,566), depression (n=1,035,760), broad depression phenotype (n=500,199) and neuroticism (n=329,821).

**Results:** We found little evidence of causal effects in either direction, except for weak evidence suggesting that UUI may reduce the risk of depression (causal odds ratio (OR) for depression: 0.99, 95% confidence interval (95%CI) 0.98-1.00; OR for broad depression 0.98, 95%CI 0.96-1.00). However, the direction of effect estimates did not consistently align across sensitivity analysis methods and the magnitude of this effect could be considered negligible.

**Conclusion:** Our study provided little evidence for causal bidirectional relationships between UI and anxiety, depression and neuroticism. This study highlights the need for further GWAS of UI (including different subtypes) with larger sample sizes.

**Highlights:**

- There was little evidence for causal bidirectional relationships between urinary incontinence (UI) and anxiety, depression, and neuroticism.
- Study limitations, including low statistical power, UI phenotype definition, and genetic instrument strength, need to be considered when interpreting the findings.
- Larger and more comprehensive GWAS of UI (including subtypes) are needed before ruling out causal relationships between UI and mental health.

## Introduction

Urinary incontinence (UI) is defined as the involuntary leakage of urine^[1]^ and is associated with adverse impacts on mental health and quality of life^[2-5]^. UI is more prevalent in women than in men^[5, 6]^, with the UK National Health Service reporting that 34% of women experience UI^[7]^. UI can be classified into three subtypes: stress urinary incontinence (SUI), characterised by urine leakage due to physical exertion, coughing or sneezing; urgency urinary incontinence (UUI), marked by leakage following a sudden, compelling need to void; and mixed urinary incontinence (MUI), which involves symptoms of both SUI and UUI^[8]^. SUI is the most common subtype (10-39%), followed by MUI (7.5-25%), and UUI (1-7%)^[9]^. Variance in prevalence estimates is attributed to differences in UI definitions and sample characteristics including age, ethnicity, and parity.

The comorbidity between UI and affective disorders, including anxiety and depression, is well established^[10-17]^, but the precise nature of this relationship is unknown. Much of the research examining this relationship is cross-sectional, limiting the ability to determine causality. Prospective studies suggest bidirectional associations between UI and anxiety/depression in women^[10]^, as well as differential associations by UI subtype^[10, 18]^. For example, a 10-year longitudinal study of 16,263 women (mean age 47 years) found that anxiety and depression increased the risk of developing both UUI and SUI in a dose-dependent manner^[10]^. In the same study, both UUI and SUI were associated with an increased incidence of mild depression and anxiety, although associations with moderate/severe cases had confidence intervals that crossed the null^[10]^. Another longitudinal study of 12,568 women (median age 58 years) found that baseline UUI was associated with a higher incidence of anxiety and depression at one year follow-up. Additionally, anxiety at baseline predicted incident UUI, whereas depression did not. The study found no evidence of bidirectional relationships between SUI and anxiety or depression^[19]^. A study of 5,291 women found evidence of prospective associations between depression and subsequent MUI, UUI, and any UI, while only SUI was associated with later depression.^[16]^ Previous studies have also found that neuroticism (a personality trait associated with an increased risk of affective disorders) is associated with an increased risk of UI^[20, 21]^.

Observational studies are prone to residual and unmeasured confounding, limiting causal inference. It is therefore unclear whether the reported relationships between anxiety/depression/neuroticism and UI reflect causal effects. Exploring the evidence for causality (unidirectional or bidirectional) between anxiety/depression/neuroticism and UI is important due to its implications for treatment strategies. If anxiety and/or depression contribute to UI risk, treatments for these disorders could be effective in alleviating UI. Conversely, if UI increases anxiety and/or depression risk, effective UI management could play a role in preventing mental illness.

This study uses Mendelian Randomization (MR) analysis^[22-24]^ to assess potential causal bidirectional relationships between anxiety/depression/neuroticism and UI in women. MR analysis uses genetic variation as a natural experiment to strengthen causal inference in observational data^[25]^. Genetic variants are used as instrumental variables for an exposure and can improve causal inference given certain core conditions: (i) the genetic instrument(s) used to proxy the exposure is robustly associated with the exposure (the relevance condition); (ii) the genetic instrument(s) used to proxy the exposure is not associated with confounders of the exposure-outcome association (independence condition); and (iii) the genetic instrument(s) used to proxy the exposure is only associated with the outcome through the exposure (exclusion-restriction condition)^[26, 27]^. Bidirectional MR can improve causal inference by independently testing the causal effects in both directions using single nucleotide polymorphisms (SNPs) identified in genome-wide association studies (GWAS).

Using GWAS summary statistics, this study conducts a two-sample MR analysis to explore bidirectional causal relationships between UI (including SUI, UUI & MUI) and anxiety, depression, and neuroticism in women. Based on existing prospective data, we hypothesise causal effects in both directions.

## Methods

### Study populations

#### Urinary incontinence GWAS

GWAS summary statistics were obtained for UI in women of European ancestry, adjusting for the first 4 principal components^[28]^. The GWAS was conducted in the Nurses’ Health Study and the Nurses’ Health Study II participant samples. These two studies employed biennial questionnaires where participants were asked about urine leakage frequency and volume. Participants responded to two UI questions: 1) “During the past 12 months, how often have you leaked urine or lost control of your urine?” with response options ranging from never to almost every day, and 2) “When you lose urine, how much usually leaks?” with response options of: a few drops, enough to wet your underwear, enough to wet outer clothing, and enough to wet the floor. These questions were asked seven times over sixteen years in the Nurses’ Health Study, and five times over 12 years in the Nurses’ Health Study II.

‘Any UI’ cases (n=6,120) were defined as individuals reporting at least weekly UI on the majority of the questionnaires (≥4 in the Nurses’ Health Study and ≥3 in the Nurses’ Health Study II), while controls (n=4,811) were those who reported never experiencing UI or experienced leaking only a few drops less than once a month on all answered questionnaires; all eligible participants must have responded to the majority of questionnaires.

Participants meeting the ‘any UI’ definition were classified into UI subtypes defined as: SUI (leaking episodes related to physical activity, coughing, sneezing, n=1,809); UUI (leaking episodes associated with a sense of urgency, n=1,942); and MUI (leaking episodes equally attributable to both activity and urgency, n=2,036). The subtype questions were not included on the questionnaires until a later stage in the study, resulting in a smaller subset of participants with subtype information compared with the total ‘any UI’ cases.

#### Anxiety, depression, and neuroticism GWAS

GWAS summary results for anxiety (n=83,566)^[29]^, depression (n=1,035,760)^[30]^, broad depression phenotype (n=500,199)^[31]^ and neuroticism (n=329,821)^[32]^ were used. The GWAS for anxiety, depression and broad depression used case status, while the neuroticism GWAS used a continuous score. All GWAS were adjusted for age, sex, and principal components and restricted to individuals of European ancestry. There was no sample overlap between the UI and anxiety, depression, and neuroticism GWAS.

The depression GWAS^[30]^ was derived from a meta-analysis of cohorts, primarily using depression cases identified via ICD-10 codes (F32-F33), though it also included depression cases based on self-report. The broad depression phenotype GWAS^[31]^ was a subset of the depression GWAS^[30]^ meta-analysis. We present results for both the depression and broad depression phenotype GWAS to explore potential differences in findings.

#### Identifying genetic instruments for exposure phenotypes

Genetic instruments were selected from the GWAS data, comprising SNPs that achieved genome-wide significance (p < 5×10^-08^). To ensure their independence for linkage disequilibrium (LD), SNPs were clumped (r = 0.001 within 10000kb) using the ieugwasr R package^[33]^ .In instances where exposure SNPs were not available within the outcome datasets, proxy variants in high LD (r^2^ > 0.8) were identified where possible, using the LDlinkR R package^[34]^.

For ‘any UI’, two independent SNPs reached genome-wide significance. No SNPs reached genome-wide significance for the UI subtypes SUI, UUI or MUI. Independent SNPs achieving genome-wide significance were identified for anxiety (5 SNPs), depression (133 SNPs), broad depression phenotype (50 SNPs) and neuroticism (75 SNPs).

For exposures with 5 or fewer SNPs reaching genome-wide significance, a sensitivity analysis using a lenient p-value threshold of 5×10^-06^ was run to expand the number of SNPs associated with the exposures. Independent SNPs reaching this threshold were identified for ‘any UI’ (23 SNPs), SUI (6 SNPs), UUI (13 SNPs), MUI (14 SNPs) and anxiety (35 SNPs.)

## Statistical analyses

### Two-sample Mendelian randomization analyses

Bidirectional MR analyses were performed between UI and its subtypes and anxiety, depression, broad depression phenotype and neuroticism. We performed harmonisation of the direction of effects between exposure and outcome associations, where non-inferable SNPs (palindromic SNPs with minor allele frequency [MAF] >0.42) were removed from the analysis. We used an inverse-variance weighted (IVW) method to meta-analyse the SNP-specific Wald ratio estimates using random effects and adjusting for heterogeneity^[35]^ to estimate the causal effect of UI on anxiety, depression and neuroticism, and vice-versa.

### Sensitivity analyses

For analyses where exposure SNPs were identified using a lenient p-value threshold of 5×10^-06^, a debiased IVW method^[36]^ was used. This estimator is built on the IVW method but incudes a bias correction factor, making it robust to the presence of weak instruments.

In the absence of horizontal pleiotropy, or when horizontal pleiotropy is balanced, the IVW estimate is unbiased^[35]^. To address potential biases stemming from SNPs acting through horizontally pleiotropic pathways, thereby violating the exclusion-restriction MR condition, we employed three additional sensitivity analysis methods. These methods, each relying on different assumptions and therefore more robust to pleiotropy, included MR-Egger regression^[37]^, weighted median ^[38]^ and weighted mode-based methods^[39]^. Results were compared across methods with a consistent effect across multiple methods strengthening causal evidence. SNP heterogeneity was assessed using Cochran’s Q statistic^[40]^.

Analyses were performed using the *TwoSampleMR R* package^[41]^ (version 0.5.6), the ieugwasr R package^[33]^ (version 0.1.5) and the *MendelianRandomization* R package (version 0.9.0). All analyses were performed using R (version 4.3.0) (www.r-project.org).

## Results

### Causal effect of urinary incontinence on anxiety/depression/neuroticism

The total number of SNPs used in each analysis after data harmonisation is presented in Table 1. Two SNPs were identified as instrumental variables for ‘any UI’, both with F-statistics greater than 31 (Supplementary table 1). There was no evidence that ‘any UI’ has a causal effect on the risk of anxiety, depression, or neuroticism (Table 2, Figure 1), with all effect estimates crossing the null. Due to the limited number of SNPs available for ‘any UI’ (two), the sensitivity analysis methods - MR-Egger regression, the weighted median method and the weighted mode-based method – were not applicable, as they require a greater number of SNPs.

**Table 1.** Number of SNPs used as exposure for each GWAS in the main MR analyses. Exposure SNPs selected using p-value threshold of 5×10^-80^.

| Exposure GWAS | Outcome GWAS | Number of SNPs in exposure GWAS | Number of exposure SNPs in outcome GWAS | Number of exposure SNPs missing from outcome GWAS | Number of missing SNPs there are proxies for | Total non-proxy and proxy SNPs used as exposure | Total SNPs remaining following harmonisation<sup>a</sup> |
| --- | --- | --- | --- | --- | --- | --- | --- |
| **(any) Urinary incontinence** | Depression | 2 | 2 | 0 | NA | 2 | 2 |
|  | Broad depression phenotype | 2 | 2 | 0 | NA | 2 | 2 |
|  | Anxiety | 2 | 2 | 0 | NA | 2 | 2 |
|  | Neuroticism | 2 | 2 | 0 | NA | 2 | 2 |
| **Depression** | (any) Urinary incontinence | 133 | 132 | 1 | 0 | 132 | 128 |
|  | Stress urinary incontinence | 133 | 130 | 3 | 0 | 130 | 126 |
|  | Urgency urinary incontinence | 133 | 128 | 5 | 0 | 128 | 125 |
|  | Mixed urinary incontinence | 133 | 130 | 3 | 0 | 130 | 126 |
| **Broad depression phenotype** | (any) Urinary incontinence | 50 | 50 | 0 | NA | 50 | 47 |
|  | Stress urinary incontinence | 50 | 49 | 1 | 0 | 49 | 46 |
|  | Urgency urinary incontinence | 50 | 49 | 1 | 0 | 49 | 46 |
|  | Mixed urinary incontinence | 50 | 49 | 1 | 0 | 49 | 46 |
| **Anxiety** | (any) Urinary incontinence | 5 | 5 | 0 | NA | 5 | 5 |
|  | Stress urinary incontinence | 5 | 5 | 0 | NA | 5 | 5 |
|  | Urgency urinary incontinence | 5 | 5 | 0 | NA | 5 | 5 |
|  | Mixed urinary incontinence | 5 | 5 | 0 | NA | 5 | 5 |
| **Neuroticism** | (any) Urinary incontinence | 75 | 75 | 0 | NA | 75 | 74 |
|  | Stress urinary incontinence | 75 | 75 | 0 | NA | 75 | 74 |
|  | Urgency urinary incontinence | 75 | 75 | 0 | NA | 75 | 74 |
|  | Mixed urinary incontinence | 75 | 75 | 0 | NA | 75 | 74 |

<sup>a</sup> = SNPs removed due to being palindromic and not able to determine forward strand using allele frequencies during the harmonisation process

**Table 2.** Bi-directional MR analyses estimates between UI phenotypes and anxiety, depression, broad depression phenotype and neuroticism. Exposure SNPs selected using p-value threshold of 5×10^-80^ .

| Exposure | Outcome | N SNPs | Causal estimate (95% CI) | P-value | Heterogeneity P-value |
| --- | --- | --- | --- | --- | --- |
| **(any) Urinary incontinence** | Anxiety | 2 | 1.05 (0.96-1.14)<sup>a</sup> | 0.26 | 0.27 |
|  | Depression | 2 | 1.02 (0.99-1.04)<sup>a</sup> | 0.27 | 0.82 |
|  | Broad depression phenotype | 2 | 1.02 (0.98-1.06)<sup>a</sup> | 0.26 | 0.76 |
|  | Neuroticism | 2 | 0.0158 (-0.0037-0.0354)<sup>b</sup> | 0.11 | 0.73 |
| **Anxiety** | (any) Urinary incontinence | 5 | 1.01 (0.82-1.26)<sup>a</sup> | 0.89 | 0.90 |
|  | Stress urinary incontinence | 5 | 1.25 (0.90-1.74)<sup>a</sup> | 0.19 | 0.31 |
|  | Urgency urinary incontinence | 5 | 0.95 (0.70-1.28)<sup>a</sup> | 0.74 | 0.81 |
|  | Mixed urinary incontinence | 5 | 0.93 (0.62-1.40)<sup>a</sup> | 0.72 | 0.10 |
| **Depression** | (any) Urinary incontinence | 128 | 0.87 (0.65-1.15)<sup>a</sup> | 0.33 | 0.003 |
|  | Stress urinary incontinence | 126 | 0.83 (0.57-1.21)<sup>a</sup> | 0.33 | 0.13 |
|  | Urgency urinary incontinence | 125 | 0.84 (0.56-1.25)<sup>a</sup> | 0.39 | 0.009 |
|  | Mixed urinary incontinence | 126 | 0.92 (0.64-1.34)<sup>a</sup> | 0.67 | 0.05 |
| **Broad depression phenotype** | (any) Urinary incontinence | 47 | 1.19 (0.85-1.67)<sup>a</sup> | 0.30 | 0.14 |
|  | Stress urinary incontinence | 46 | 1.22 (0.78-1.92)<sup>a</sup> | 0.39 | 0.35 |
|  | Urgency urinary incontinence | 46 | 1.13 (0.73-1.77)<sup>a</sup> | 0.58 | 0.34 |
|  | Mixed urinary incontinence | 46 | 1.04 (0.62-1.74)<sup>a</sup> | 0.88 | 0.02 |
| **Neuroticism** | (any) Urinary incontinence | 74 | 1.08 (0.88-1.32)<sup>a</sup> | 0.48 | 0.43 |
|  | Stress urinary incontinence | 74 | 0.99 (0.73-1.36)<sup>a</sup> | 0.97 | 0.13 |
|  | Urgency urinary incontinence | 74 | 1.06 (0.80-1.41)<sup>a</sup> | 0.67 | 0.96 |
|  | Mixed urinary incontinence | 74 | 1.16 (0.84-1.59)<sup>a</sup> | 0.37 | 0.04 |

**Figure 1.**
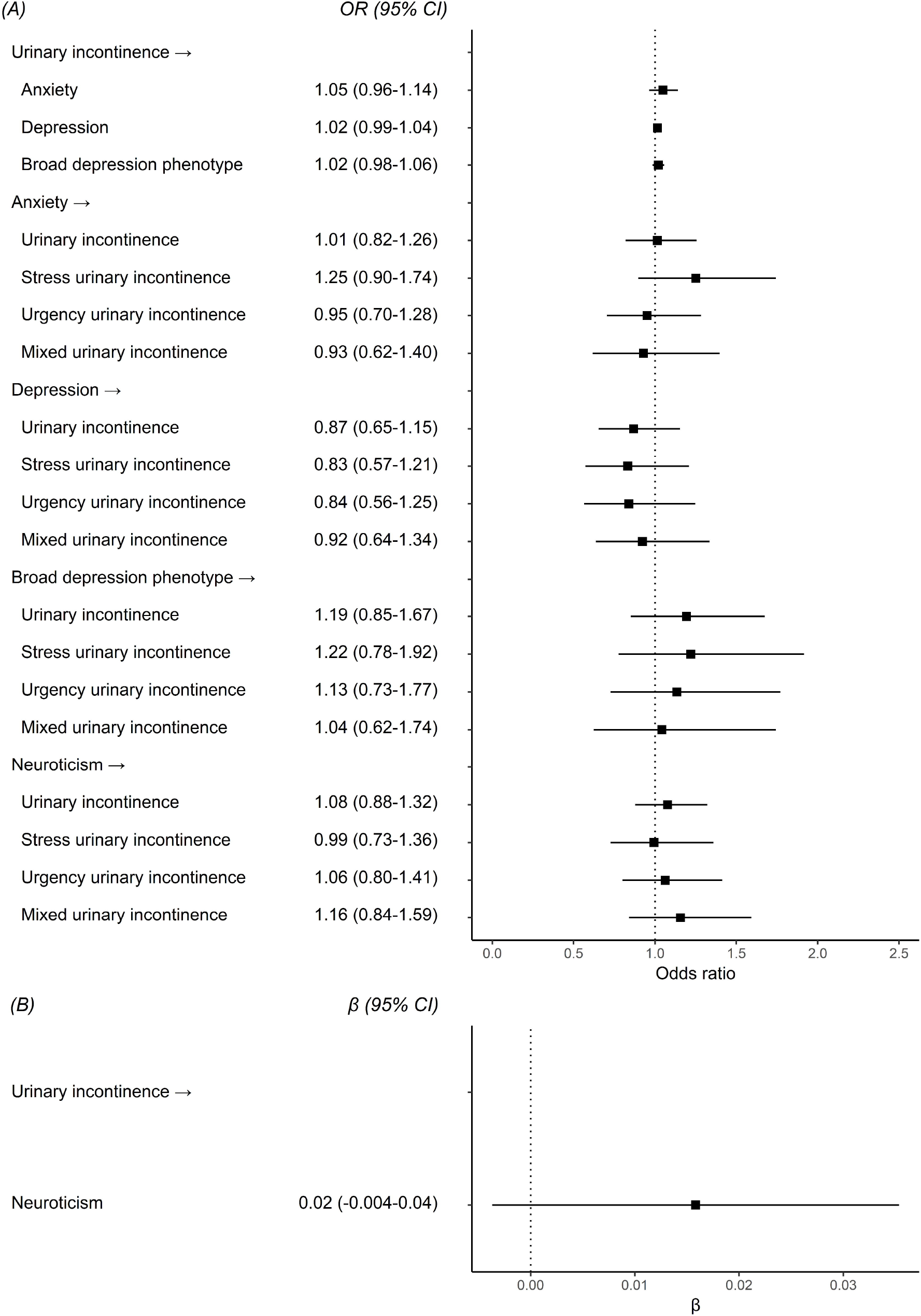
Bi-directional MR analyses estimates between UI phenotypes and anxiety, depression, broad depression phenotype and neuroticism. All estimates are IVW MR analyses. Exposure SNPs selected using p-value threshold of 5×10^-80^. (A) MR estimates represent the odds ratio (OR) and 95% CI for an approximate doubling of the genetic liability to presence (vs absence) of each binary exposure. (B) MR estimates represent estimated change (beta) in total neuroticism score for an approximate doubling of the genetic liability to presence (vs absence) of each binary exposure.

To explore causal relationships across all UI subtypes (‘any UI’, SUI, UUI, MUI), we applied a lenient p-value threshold to identify additional SNPs to instrument the exposures. The total number of SNPs used in each analysis after data harmonisation is presented in Supplementary Table 2. There was little evidence that any of the UI subtypes (‘any UI’, SUI, UUI, MUI) had a causal effect on anxiety, depression or neuroticism (Table 3, Figure 2). Contrary to our hypothesis, genetic liability for UUI was associated with a decreased risk of depression (OR=0.987; 95% CI: 0.975-0.998; P-value=0.02) and the broad depression phenotype (OR=0.980; 95% CI: 0.963-0.997; P-value=0.02). However, the direction of effect estimates did not consistently align across sensitivity analysis methods (Supplementary Table 3) and the magnitude of this effect could be considered negligible.

**Table 3.** Bi-directional MR analyses estimates between UI phenotypes and anxiety, depression, broad depression phenotype and neuroticism. Exposure SNPs selected using a lenient p-value threshold of 5×10^-80^.

| Exposure | Outcome | N SNPs | Causal estimate (95% CI) | P-value |
| --- | --- | --- | --- | --- |
| **(any) Urinary incontinence** | Anxiety | 21 | 1.02 (0.99-1.04)<sup>a</sup> | 0.15 |
|  | Depression | 22 | 1.01 (1.00-1.02)<sup>a</sup> | 0.27 |
|  | Broad depression phenotype | 21 | 1.01 (1.00-1.02)<sup>a</sup> | 0.22 |
|  | Neuroticism | 22 | 0.0061 (-0.0005-0.0127)<sup>b</sup> | 0.07 |
| **Stress urinary incontinence** | Anxiety | 6 | 1.01 (0.96-1.06)<sup>a</sup> | 0.80 |
|  | Depression | 6 | 1.00 (0.98-1.01)<sup>a</sup> | 0.72 |
|  | Broad depression phenotype | 4 | 1.00 (0.98-1.01)<sup>a</sup> | 0.56 |
|  | Neuroticism | 6 | -0.008 (-0.027-0.012)<sup>b</sup> | 0.44 |
| **Urgency urinary incontinence** | Anxiety | 11 | 0.98 (0.94-1.02)<sup>a</sup> | 0.41 |
|  | Depression | 10 | 0.99 (0.98-1.00)<sup>a</sup> | 0.02 |
|  | Broad depression phenotype | 10 | 0.98 (0.96-1.00)<sup>a</sup> | 0.02 |
|  | Neuroticism | 10 | -0.006 (-0.022-0.011)<sup>b</sup> | 0.50 |
| **Mixed urinary incontinence** | Anxiety | 13 | 1.00 (0.96-1.05)<sup>a</sup> | 0.99 |
|  | Depression | 13 | 1.00 (0.99-1.01)<sup>a</sup> | 0.66 |
|  | Broad depression phenotype | 13 | 1.01 (0.99-1.02)<sup>a</sup> | 0.35 |
|  | Neuroticism | 13 | 0.0027 (-0.0141-0.0195)<sup>b</sup> | 0.75 |
| **Anxiety** | (any) Urinary incontinence | 35 | 0.99 (0.87-1.13)<sup>a</sup> | 0.87 |
|  | Stress urinary incontinence | 28 | 1.01 (0.84-1.21)<sup>a</sup> | 0.93 |
|  | Urgency urinary incontinence | 27 | 0.93 (0.77-1.13)<sup>a</sup> | 0.47 |
|  | Mixed urinary incontinence | 27 | 1.06 (0.86-1.32)<sup>a</sup> | 0.57 |

**Figure 2.**
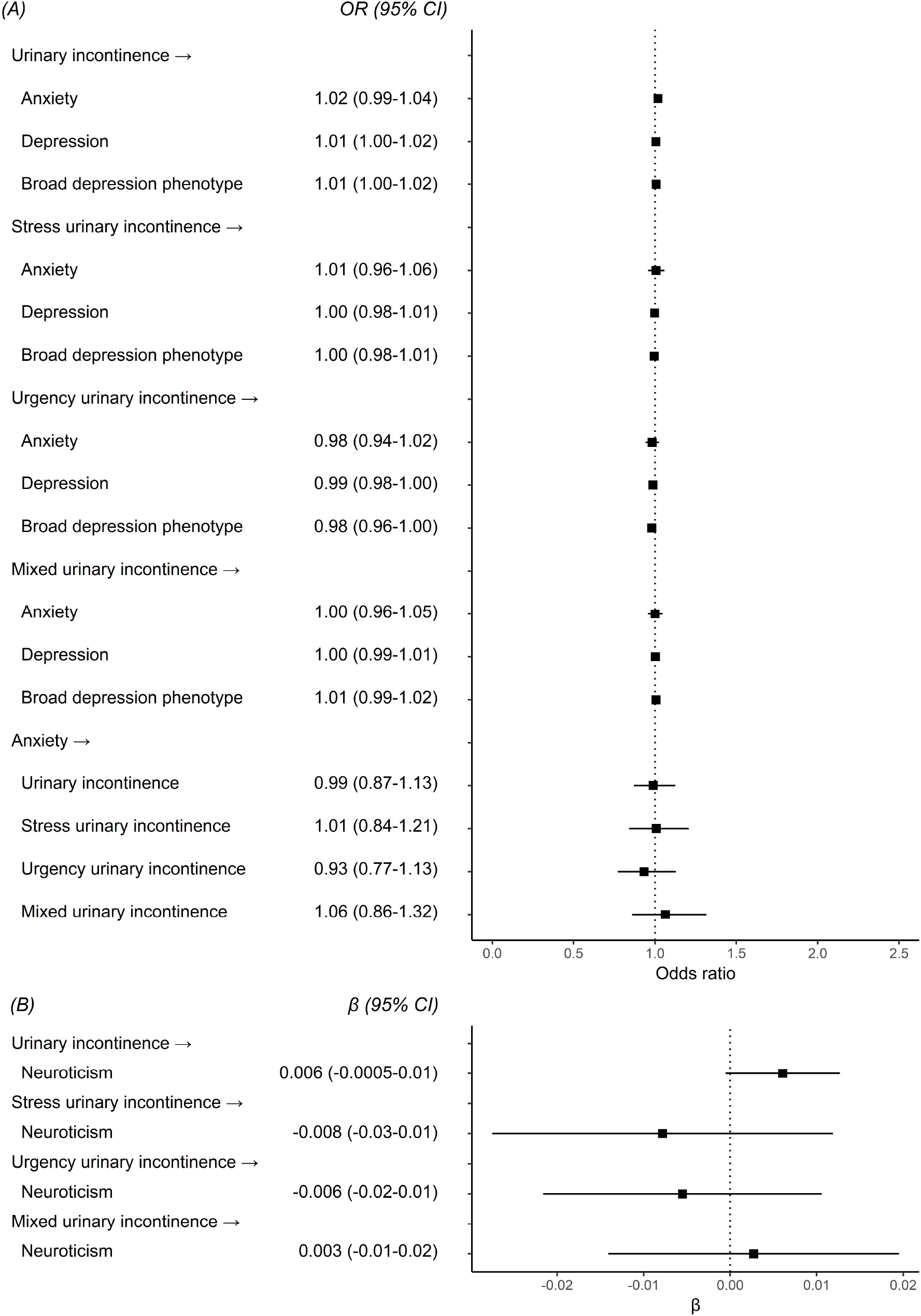
Bi-directional MR analyses estimates between UI phenotypes and anxiety, depression, broad depression phenotype and neuroticism. All estimates are IVW MR analyses. Exposure SNPs selected using a lenient p-value threshold of 5×10^-80^. (A) MR estimates represent the odds ratio (OR) and 95% CI for an approximate doubling of the genetic liability to presence (vs absence) of each binary exposure. (B) MR estimates represent estimated change (beta) in total neuroticism score for an approximate doubling of the genetic liability to presence (vs absence) of each binary exposure.

### Causal effect of anxiety/depression/neuroticism on urinary incontinence

The total number of SNPs used in each analysis after data harmonisation is summarised in Table 1, with all F-statistics exceeding 29 (Supplementary table 1). There was little evidence for causal effects of anxiety, depression or neuroticism on any of the UI subtypes (‘any UI’, SUI, UUI or MUI), with all confidence intervals crossing the null (Table 2, Figure 1).

Results from sensitivity methods generally showed directional inconsistencies with the IVW estimates, with most confidence intervals crossing the null (Supplementary Table 4). In MR-Egger analyses, genetic liability for the broad depression phenotype was associated with an increased risk of ‘any UI’ (OR=8.85; 95% CI: 1.40-56.17; P-value 0.03) and MUI (OR=14.94; 95% CI: 1.04-214.13; P-value 0.05) (Supplementary Table 4). However, given the large effect sizes, wide confidence intervals and inconsistencies with the IVW estimates, these findings should be interpreted with caution. Such large estimates may indicate potential bias, including horizontal pleiotropy or weak instrument bias, which can inflate effect sizes and reduce precision in MR-Egger^[38, 40]^. The wide confidence intervals further suggest limited precision, likely due to overfitting or insufficient statistical power, particularly given the small sample size of the UI GWAS^[42, 43]^.

Given that only 5 SNPs were identified as genetic instruments for anxiety, a lenient p-value threshold was used to identify additional SNPs, increasing the number of instruments for anxiety to 35 (Supplementary 2). This addition of SNPs had little effect upon the effect estimates when compared to the main analysis (Table 3, Figure 2, Supplementary Table 4).

## Discussion

We used bidirectional two-sample MR analysis to investigate the causal relationships between UI and anxiety, depression and neuroticism. We found little evidence of causal effects in either direction, except for weak evidence from sensitivity analyses suggesting that genetic liability for UUI may reduce the risk of depression - contrary to our initial hypothesis. Additionally, the sensitivity analysis method MR-Egger, indicated that the broad depression phenotype might increase the risk of ‘any UI’ and MUI, though this result should be interpreted with caution.

To our knowledge, only one previous study has used two-sample MR to examine the influence of depressive symptoms, anxiety and neuroticism on urinary frequency/incontinence^[44]^. This study reported a minimal association between neuroticism and urinary frequency/incontinence (OR: 1.001; 95% CI:1.000–1.001; P-value < 0.01), although the effect size was negligible. While that study had larger GWAS sample sizes for both the exposure (n= 374,323) and the outcome (n= 462,933), the outcome GWAS had limitations. Specifically, the prevalence of UI was low and likely underestimated (1,624 cases among 461,309 controls), and the definition of UI was less specific, relying on non-standardized self-reported leakage rather than a validated questionnaire. These methodological differences may help explain some of the discrepancies in findings between their findings and ours.

While we found a weak association between UUI and reduced risk of depression, we are cautious in interpreting these results, given several factors, including multiple testing, the lenient p-value threshold for SNP selection, and the small effect sizes observed. MR-Egger sensitivity analysis also suggested a potential association between broad depression and increased risk of UI and MUI. However, these estimates were large, with wide confidence intervals wide, and inconsistent with the IVW estimates. It is important to note that MR-Egger estimates can be biased if the MR InSIDE assumption is violated^[43, 45]^, and the large effect sizes may reflect such violations, such as horizontal pleiotropy or weak instrument bias. Given the lack of evidence from the IVW method, these sensitivity findings should be interpreted with caution.

Our findings are inconsistent with previous observational studies, which have reported prospective bidirectional associations between UI and depression/anxiety.^[10, 19]^ Other studies have also found neuroticism to be associated with an increased risk of UI.^[20, 21]^ These discrepancies may suggest that the associations reported in observational studies are influenced by confounding factors. Co-twin control studies have suggested that familial factors, such as intrauterine exposures, maternal factors, and early-life environment, may confound the relationship depressive symptoms and UI subtypes^[20]^. Another explanation for the lack of causal bidirectional effects in our study is low statistical power. While low statistical power – particularly when examining UI as the exposure due to the small sample size – may have contributed to the null findings. In the reverse direction, we had greater power but still there was little evidence of a causal effect of mental health on incontinence risk.

A key limitation of this study is the limited number of SNPs robustly associated with UI, which reduced the strength of our genetic instruments. This limitation is largely due to the relatively small sample size of the UI GWAS. Additionally, the self-reported nature of the UI phenotypes may have led to misclassification of UI cases, affecting instrument selection and MR estimates. While a larger and more recent UI GWAS (n= 13,066 women)^[18]^ was available, full genome-wide summary statistics necessary for bi-directional MR analyses were only available for a subset of women (n=8,997). Additionally, this GWAS adjusted for BMI and parity, which are known risk factors for UI. Adjusted GWAS summary statistics may introduce bias in MR analyses in ways that are difficult to predict and that are highly dependent on the underlying (and unobserved) causal structure.^[46]^ This highlights the need for larger-scale GWAS of urinary incontinence that are unadjusted for UI risk factors.

Another limitation relates to the selection of participants. Given the predominance of White European samples in publicly available GWAS data, our findings may lack generalizability across diverse populations. Conducting more diverse GWAS encompassing a broader range of ethnicities and age groups will significantly enhance the transferability and external validity of the findings.

A final limitation concerns the inclusion of both males and females in the depression, anxiety and neuroticism GWAS, while the UI GWAS included females only. A core condition for valid two-sample MR is that the exposure and outcome GWAS are conducted in the same population^[47]^. Although we have assumed this condition held, despite the UI GWAS only being performed in women, any violation of this assumption could introduce bias, as genetic associations with the outcome may not be reproduced in the exposure population^[48]^. Therefore, causal effects between UI and depression/anxiety/neuroticism may not have been identified. This additionally highlights the need for large-scale sex-specific GWAS especially for outcomes where sex-specific genetic factors may play an important role in outcome susceptibility.

Given these limitations, further research with larger samples for genetic discovery, coupled with more refined phenotypes for UI (including UI subtypes) is needed.

## Conclusion

Our study provided little evidence for causal bidirectional relationships between UI and anxiety, depression and neuroticism. To enable well-powered MR studies, there is a need for further GWAS of UI (including different subtypes) with larger sample sizes. Additionally, more diverse GWAS that include individuals from different ethnic backgrounds and sex-specific analyses will be crucial for improving the generalizability of findings.

## Data availability

Publicly available GWAS summary statistics have been used in this paper. Information on how to obtain/request this data is available in Supplementary table 5.

All code used to run analyses is available on GitHub: https://github.com/RochelleKnight/urinary-incontinence-and-mental-health

## Author contributions

CJ, AF and AGS conceived the study. CJ, AF, AGS, TP, KB and RK contributed to its design and methodology. RC provided clinical direction and interpretation of results. RK conducted statistical analyses. RK produced the first draft of the manuscript. All authors critically appraised the manuscript for important intellectual content and contributed to its final version.

